# Nasal high-frequency percussive ventilation versus nasal continuous positive airway pressure in term and preterm neonates exhibiting respiratory distress: a randomized controlled trial (TONIPEP; NCT 02030691)

**DOI:** 10.1101/2020.02.11.20022178

**Authors:** Renesme Laurent, Dumas de la Roque Eric, Germain Christine, Chevrier Agnès, Rebola Muriel, Cramaregeas Sophie, Benard Antoine, Elleau Christophe, Tandonnet Olivier

## Abstract

**Objective:** To determine whether the use of nasal, high-frequency percussive ventilation (nHFPV) to manage neonatal respiratory distress decreases the regional cerebral oxygen saturation (rScO_2_) below that afforded by nasal continuous positive airway pressure (nCPAP).

**Design:** Monocentric, prospective, randomized, monocentric, open-label, non-inferiority crossover trial.

**Patients:** Newborns of gestational age (GA) ≥ 33 weeks exhibiting persistent respiratory distress after 10 min of life (Silverman score ≥ 4).

**Intervention:** nHFPV and nCPAP, in succession and in random order.

**Main outcome measure:** Mean rScO_2_, as revealed by near-infrared spectroscopy (NIRS) performed over the last 5 min of each ventilation mode. To show that nHFPV was not inferior to nCPAP, our *a priori* calculations required that the lower boundary of the bilateral 95% confidence interval (CI) of the difference between the mean rScO_2_ values of each ventilation mode should exceed –5.

**Results:** Forty-nine newborns were randomized and 46 were analyzed. The mean (± standard deviation [SD]) GA and birth weight were 36.4 ± 1.9 weeks and 2,718 ± 497 g. The diagnosis was transient tachypnea in 65% of cases and respiratory distress syndrome in 35%. The mean rScO_2_ difference during the last 5 min of each ventilation mode (nHFPV minus nCPAP) was – 0.7 ± 5.4% (95% CI –2.25; 0.95). Neither a period effect nor a period-treatment interaction was evident. The mean transcutaneous carbon dioxide values (n = 26) for nCPAP and nHFPV were 7.1 ± 4.8 and 7.9 ± 5.1 kPa, respectively. No harmful or unintentional effect was observed.

**Conclusion:** In our study on newborns of GA ≥ 33 weeks treated for respiratory distress, cerebral oxygenation via nHFPV was not inferior to nCPAP.

**What is already known on the topic:**

- Non-invasive high-frequency ventilation is feasible in preterm newborns and seems to improve ventilation compared to nasal CPAP.
- We previously showed that nasal high-frequency percussive ventilation (nHFPV) was more efficient that nCPAP for respiratory distress management in newborns of gestational age (GA) ≥ 35 weeks.
- The impact of mechanical ventilation, especially high-frequency modes, on cerebral blood flow in neonates is of concern.

**What this study adds:**

- nHFPV was well-tolerated and non-inferior to nasal CPAP as measured by rScO_2_ levels when used to manage respiratory distress at birth in newborns of GA ≥ 33 weeks.

## Introduction

Pulmonary respiratory distress is the leading cause of neonatal morbidity and mortality regardless of the term status [1]. The overall incidence is 7% of live births, and the incidence varies with the gestational age [1,2]. Neonatal respiratory distress is a heterogeneous entity, with different etiologies exhibiting variable evolutions. The main cause of respiratory distress is transient tachypnea of the newborn (TTN) (> 40% of all respiratory distress) [3]. Other causes include respiratory distress syndrome (RDS), meconium aspiration syndrome, infection, persistent pulmonary hypertension, and pneumothorax [4]. Initial management involves establishment of non-invasive respiratory support to improve oxygenation and reduce breathing effort. Nasal continuous positive airway pressure (nCPAP) is the most widely used noninvasive respiratory support for neonates [5]. High-frequency percussive ventilation (HFPV) is a pressure-limited, time-cycled high-frequency mode of ventilation that delivers small bursts of high-flow respiratory gas at subphysiological tidal volumes 90 to 650 times per min. HFPV may be invasive or non-invasive [6]. The VDR3c (Percussionaire^®^ Corporation) is a pneumatically powered, time-cycled pressure-limited ventilator featuring a sliding venturi system (Phasitron, Percussionaire^®^ Corporation) powered by compressed gas. We previously showed that nHFPV was more efficient that nCPAP for respiratory distress management in newborns of gestational age (GA) ≥ 35 weeks [7].

Our objective here was to compare the tolerance of nHFPV and nCPAP used to manage neonatal respiratory distress in newborns of GA ≥ 33 weeks. Tolerance was assessed by measuring regional cerebral oxygen saturation using near-infrared spectroscopy (NIRS).

## Methods

### Study design and settings

We performed a prospective, randomized, open-label, two-period, crossover trial. We conducted intra-individual comparisons of the two ventilation strategies; each newborn served as his/her own control. The study was performed at the University Hospital of Bordeaux (a tertiary center) from May 2014 to November 2016. The trial was approved by our institutional review board (Comité de Protection des Personnes Sud-Ouest et Outre Mer III) and was registered in the NIH clinical trials registry (registration number [clinicaltrials.gov]: NCT02030691).

### Participants

Inclusion criteria were birth (vaginal delivery or C-section) at the University Hospital of Bordeaux, GA ≥ 33 weeks, birth weight ≥ 1,000 g, and respiratory distress (Silverman score ≥ 4) evident after 10 min of life. Newborns with meconium aspiration syndrome or congenital malformations were excluded.

### Randomization

A randomization list was prepared prior to the study by a statistician from our Clinical Epidemiology Unit. Randomization was balanced (1:1 ratio) between the two groups; the block size was 6, and no factor was stratified. After parental consent was obtained, randomization was performed using the secure website of the Clinical Epidemiology Unit.

### Interventions

Newborns in respiratory distress (Silverman score ≥ 4 after 10 min of life) were admitted to the neonatal intensive care unit and initially received nCPAP. After parental consent was received, and randomization performed, all newborns were allocated to either nCPAP followed by nHFPV or nHFPV followed by nCPAP. Each ventilation mode was used for 15 min (30 min in total). Non-invasive ventilation was ceased if either respiratory distress worsened to an extent requiring intubation and mechanical ventilation, or if clinical evolution allowed weaning of the support. We used a Babylog 8000-Plus (Dräger^®^) to deliver nCPAP and a VDR-3c fitted with a Monitron II pressure monitor (Percussionaire^®^ Corporation) to deliver nHFPV. Both ventilators featured heated and humidified breathing circuits and the same nasal probes.

### Primary outcome

The primary outcome was the average regional cerebral oxygen saturation (rScO_2_) during the last 5 min of each ventilation mode. rScO_2_ was non-invasively measured by NIRS (INVOS Cerebral/Somatic Oximeter; Medtronic^®^) using a probe placed on the forehead. Recording of rScO_2_ commenced at inclusion, and all investigators were blinded to the rScO_2_ data.

### Secondary outcomes

#### Transcutaneous carbon dioxide (TcPCO_2_) measurement

TcPCO_2_ (TCM TOSCA monitor; Radiometer^®^) was continuously recorded after inclusion. A capillary blood sample was taken for blood gas analysis.

### Vital parameters

The transcutaneous oxygen saturation (SpO_2_), heart rate (HR), and respiratory rate (RR) were continuously measured (Dash 2000 monitor; GE Healthcare^®^). The vital parameters (SpO_2_, HR, and RR), mean arterial blood pressure, axillary temperature, and Silverman score were collected at inclusion and every 5 min during the study.

### Ventilator settings

For nCPAP, the ventilator settings recorded were the positive end-expiratory pressure (PEEP) and the fraction of inspired oxygen (FiO_2_). For nHFPV, the mean airway pressure (MAP), percussion frequency, and FiO_2_ were noted. All settings were recorded every 5 min. If a setting required adjustment between these timepoints, the following parameters were noted: time (min), SpO_2_, HR, RR, and the new ventilator setting.

### Respiratory distress diagnosis

If respiratory distress decreased, allowing weaning of respiratory support, we diagnosed TTN. If respiratory distress worsened (as evidenced by oxygen dependence or a poorer Silverman score) to an extent requiring intubation and administration of exogenous surfactant, we diagnosed RDS.

### Statistical analysis

This non-inferiority crossover trial compared two ventilation strategies administered to all newborns in random order. The study was adequately powered; it was possible to explore the non-inferiority of nHFPV compared to nCPAP in terms of the mean rScO_2_ over the last 5 min of either modality. The literature [8,9] indicated that the mean rScO_2_ observed in such patients is approximately 80%, with a standard deviation (SD) of 10%. We defined nHFPV as being non-inferior to nCPAP if the lower boundary of the bilateral 95% CI for the difference (nHFPV minus nCPAP) was less than –5%; otherwise, the new ventilation would be poorer than the reference ventilation. Our sample size of 80 patients afforded a power of 80% in terms of revealing non-inferiority of the new regimen, assuming a non-inferiority margin of –5 (nQuery software ver. 6.0). The results are expressed as means with 95% confidence intervals (CIs) or SDs for continuous variables, and as frequencies for categorical variables. We used a linear regression model to assess the effects of treatment, treatment period (first or second), and treatment-period interactions. The latter exploration checked for the absence of a carryover effect, which would have compromised the analytical validity of the data from the second treatment period. All statistical analyses were conducted by a biostatistician from our Clinical Epidemiology Unit using SAS software ver. 9.4 (SAS Institute Inc., Cary, NC, USA). We performed intention-to-treat analyses. Primary outcome analysis featured replacement of missing rScO_2_ data by the poorest sample value. Sensitivity analyses were performed on all available data. Maximum bias analysis proceeded by replacing missing rScO_2_ data by the poorest value in one group and the best value in the other group, and vice versa.

## Results

### Study population

Forty-nine newborns were included. Inclusions were suspended in November 2016 because it became difficult to enroll new participants. Forty-six participants were analyzed (two were excluded because of consent default from one of their two parents; and one because of rapid deterioration just after inclusion, triggering a need for intubation and exogenous surfactant administration). A study flowchart is shown in Figure 1. The mean gestational age and birth weight were 36.4 ± 1.9 weeks and 2,718 ± 497 g, respectively. At inclusion, the mean SpO_2_ (± SD) was 96 ± 4%, the mean RR was 53 ± 21 cycles/min, and the mean Silverman score (without respiratory support) was 5 ± 1. The initial ventilator setting for nCPAP was PEEP +5 cm H_2_O; for nHFPV, the settings were MAP +5 cm H_2_O and a percussion frequency of 600 cycles/min. Demographic data are listed in Table 1.

**Table 1.** Demographic data.

|  | **Study population**<br>**n=46** | **nCPAP followed by**<br>**nHFPV**<br>**n=22** | **nHFPV followed by**<br>**nCPAP**<br>**n=24** |
| --- | --- | --- | --- |
| Antenatal steroids<br>n (%) | 10 (22) | 3 (14) | 7 (30) |
| C-section<br>n (%) | 22 (48) | 11 (50) | 11 (46) |
| Male<br>n (%) | 26 (56) | 12 (54) | 14 (58) |
| Gestational age<br>(weeks of gestation) | 36.4 ± 1.9 | 36.3 ± 1.9 | 36.4 ± 2 |
| Birthweight<br>(grams) | 2718 ± 497 | 2813 ± 534 | 2631 ± 455 |
| Apgar 5 min | 9 ± 1 | 9 ± 1 | 9 ± 1 |
| Capillary blood gas<br>(inclusion) |  |  |  |
| pH | 7.2 ± 0.1 | 7.3 ± 0.1 | 7.2 ± 0.1 |
| PaCO<sub>2</sub> (kPa) | 7.6 ± 2.2 | 7.3 ± 1.9 | 7.8 ± 2.5 |
| HCO<sub>3</sub><sup>-</sup> (mmol/L) | 23.3 ± 2.8 | 23.6 ± 3.5 | 23.1 ± 2.1 |
| Lactate<br>(mmol/L) | 3.5 ± 1.8 | 3.6 ± 1.8 | 3.4 ± 1.9 |

Results presented as mean ± standard deviation.
PaCO<sub>2</sub> : partial pressure of carbon dioxide ; HCO<sub>3</sub><sup>-</sup> : bicarbonate level.

**Figure 1.**
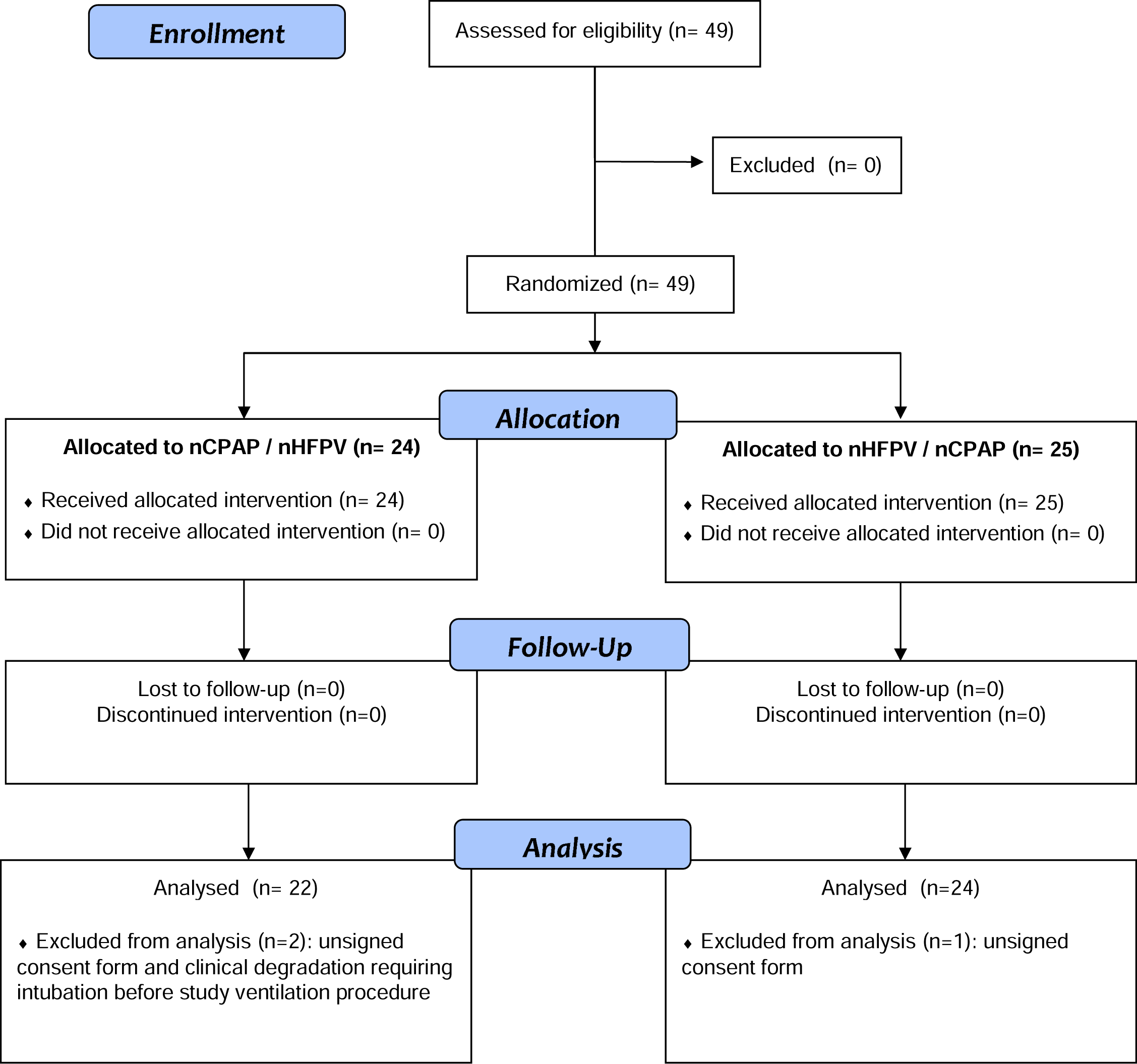
Participants flow chart. nHFPV: nasal High-Frequency Percussive Ventilation. nCPAP: nasal Continuous Positive Airway Pressure.

### Intervention

All participants (n = 46) underwent complete ventilation sequences (nCPAP followed by nHFPV or nHFPV followed by nCPAP). No unexpected or harmful effect, or death, was noted.

### Outcomes

#### Primary outcome

The mean rScO_2_ values over the last 5 min of ventilation (ten rScO_2_ measurements by patient and ventilation mode) were 82 ± 9% for the nCPAP mode and 82 ± 8% for the nHFPV mode. For four patients, at least one rScO_2_ value was missing. The mean difference of the rScO_2_ means (nHFPV minus nCPAP) was –0.7 ± 5.4% (95% CI –2.25; 0.95%). Neither a period effect nor a period-treatment interaction was found. Analysis of all available data yielded a mean rScO_2_ difference of –0.2% (95% CI –1.53; 1.21%). On maximum bias analysis, the mean rScO_2_ difference was 2.8% (95% CI –0.46; 6.11%) after replacing missing rScO_2_ data by the best observed value (95%) for nHFPV and the worst observed value (55%) for nCPAP. The difference was –3.3% (95% CI –6.58; 0.06%) when missing rScO_2_ values were replaced by the worst observed value (55%) for nHFPV and the best observed value (95%) for nCPAP.

#### Secondary outcomes

TcPCO_2_ data were available for only 26 participants; the data are listed in Table 2. The mean TcPCO_2_ levels during the last 5 min of each ventilation mode were 7.1 ± 4.8 kPa for the nCPAP mode and 7.9 ± 5.1 kPa for the nHFPV mode. Vital parameters and ventilator settings are listed in Table 3. Thirty participants (65%) were finally weaned from respiratory support and were diagnosed with TTN. Sixteen (35%) required intubation and exogenous surfactant and were diagnosed with RDS.

**Table 2.** Demographic data of participants with TcPCO_2_ measurement.

|  | **Study population**<br>**n=46** | **Participants with TcPCO<sub>2</sub>**<br>**measurement**<br>**n=26** |
| --- | --- | --- |
| Male<br>n (%) | 26 (56) | 13 (50) |
| Gestational age<br>(weeks of gestation) | 36.4 ± 1.9 | 36.4 ± 2.2 |
| Birthweight<br>(grams) | 2718 ± 497 | 2641 ± 457 |
| Apgar 5 min | 9.4 ± 0.9 | 9.4 ± 1 |
| Silverman score<br>(inclusion) | 5.3 ± 1.2 | 5.3 ± 1.4 |
| Capillary blood gas<br>(inclusion) |  |  |
| pH | 7.2 ± 0.1 | 7.3 ± 0.1 |
| PaCO<sub>2</sub> (kPa) | 7.6 ± 2.2 | 7.4 ± 2.5 |
| HCO<sub>3</sub><sup>-</sup> (mmol/L) | 23.3 ± 2.8 | 22.6 ± 2.8 |
| Lactate (mmol/L) | 3.5 ± 1.8 | 3.5 ± 1.8 |

Results presented as mean ± standard deviation.
TcPCO<sub>2</sub>: Transcutaneous carbon dioxide measurement.
PaCO<sub>2</sub>: partial pressure of carbon dioxide ; HCO<sub>3</sub><sup>-</sup>: bicarbonate level.

**Table 3.** Vital parameters and ventilator settings.

|  |  | nCPAP |  |  | nHFPV |  |  |
| --- | --- | --- | --- | --- | --- | --- | --- |
|  |  | M5 | M10 | M15 | M5 | M10 | M15 |
| Vital parameters | SpO<sub>2</sub> (%) | 97 ± 4 | 97 ± 3 | 97 ± 3 | 97 ± 4 | 97 ± 4 | 97 ± 3 |
|  | Respiratory rate (cycle/min) | 58 ± 21 | 56 ± 26 | 54 ± 22 | 54 ± 24 | 55 ± 21 | 49 ± 19 |
|  | Heart rate (beats per minute) | 144 ± 14 | 144 ± 14 | 142 ± 15 | 147 ± 16 | 147 ± 14 | 146 ± 13 |
|  | MABP (mmHg) | 40 ± 6 | 40 ± 6 | 40 ± 6 | 42 ± 6 | 41 ± 6 | 40 ± 5 |
| Ventilators settings | FiO<sub>2</sub> (%) | 23 ± 5 | 23 ± 4 | 23 ± 3 | 23 ± 5 | 23 ± 5 | 23 ± 5 |
|  | PEEP (cmH<sub>2</sub>O) | 5 ± 0.4 | 5 ± 0.4 | 5 ± 0.4 | - | - | - |
|  | MAP (cmH<sub>2</sub>O) | - | - | - | 5.5 ± 1 | 5.5 ± 0.9 | 5.6 ± 0.9 |
|  | Percussion frequency (cycle/min) | - | - | - | 604 ± 37 | 601 ± 38 | 601 ± 38 |

Results presented as mean ± standard deviation.
nCPAP : nasal Continuous Positive Airway Pressure.
nHFPV ; nasal High Frequency Percussive Ventilation.
Timepoints (M5, M10, M15) : variables were collected at 5, 10 and 15 minutes for each ventilation mode.
SpO<sub>2</sub> : Transcutaneous oxygen saturation.
MABP : Mean arterial blood pressure.
FiO<sub>2</sub> : fraction of inspired oxygen.
PEEP : Positive End-Expiratory Pressure.
MAP : Mean Airway Pressure.

## Discussion

nHFPV was well-tolerated and non-inferior to nCPAP as measured by rScO_2_ levels when used to manage respiratory distress at birth in newborns of GA ≥ 33 weeks. Sensitivity analyses confirmed this, except when replacing missing rScO_2_ values by the worst observed value under nHFPV and the best observed value under nCPAP. However, it is unlikely that all nCPAP rScO_2_ data would be compatible with the 55% value under nHFPV. Indeed, most of the missing data were attributable to signal loss (contact issues between the sensor and forehead); all other vital parameters (SpO_2_, HR, and RR) collected at the same time were within normal ranges.

If respiratory distress is evident, PEEP improves alveolar recruitment and limits alveolar collapse at the end of expiration, increasing residual functional capacity and decreasing breathing effort. PEEP also improves the ventilation/perfusion ratio and lung compliance, and reduces airway resistance [10]. A meta-analysis by Li et al. compared nasal high-frequency oscillatory ventilation (nHFOV) to nCPAP as primary respiratory support or as respiratory support after extubation in preterm infants. nHFOV was associated with better CO_2_ elimination than afforded by nCPAP and reduced the risk of intubation [11]. Effectiveness of nHFOV compared to nCPAP for the prevention of reintubation after extubation in preterm infants was confirmed by a recent large randomized controlled study [12]. Mukerji et al. showed on a term newborn mannequin that nHFOV allowed a better CO_2_ elimination than afforded by nCPAP [13].

The impact of mechanical ventilation (invasive or not) on cerebral blood flow in neonates is of concern. Such flow may be affected by positive pressure, PaCO_2_ variation (hypocapnia and cerebral vasodilatation), and, to a lesser extent, PaO_2_ variation. Ventilation with positive pressure increases intrathoracic pressure reducing the right ventricular preload and cardiac output, thus decreasing cerebral blood flow. Dani et al. showed that a PEEP of 2 to 6 cm H_2_O did not impair cerebral oxygenation or cerebral blood volume as assessed via NIRS in preterm infants of GA < 30 weeks [14]. Milan et al. have shown that in preterm infants, nCPAP (compared to invasive ventilation, thus synchronized intermittent ventilation and HFV) was associated with better cerebral blood flow [15]. To the best of our knowledge, no study has assessed cerebral oxygenation when non-invasive HFV is used to relieve neonatal respiratory distress. In our present study, the positive pressures used (PEEP and MAP) and the TcPCO_2_ data were similar for both ventilation modes (nCPAP and nHFPV).

We chose to investigate the nHFPV form of non-invasive high-frequency ventilation. HFPV can be delivered either invasively or non-invasively. As is the case for other high-frequency devices, HFPV generates a sub-physiological tidal volume (2 to 3 mL/kg) at high frequency (over 9 Hz). The HFPV system uses a sliding venturi device (Phasitron) powered by compressed gas to generate the tidal volumes. The Phasitron serves as both the inspiratory and expiratory valve [6,16]. Non-invasive HFPV enhances secretion clearance and is used to deliver respiratory physiotherapy to patients with cystic fibrosis, neuromuscular disorders, and chronic obstructive pulmonary disease [17,18]. We previously showed that use of nHFPV to manage TTN in term newborns significantly decreased oxygen requirements and the duration of respiratory distress compared to nCPAP. nHFPV was well tolerated; no adverse effect was reported [7]. We also showed, using an animal model of meconium aspiration syndrome, that invasive HFPV afforded better oxygenation and a lower MAP than HFOV [19]. The advantages of nHFPV over other high-frequency modes include better secretion clearance and use of an open breathing circuit that limits barotrauma and allows the patient to breathe spontaneously without effort. In the present study, the initial nHFPV settings (percussion frequency 10 Hz, MAP +5 cm H_2_O) were similar to those of previous studies [13,20,21].

Our crossover design limits bias caused by inter-individual variations in rScO_2_ attributable to the seriousness and cause of respiratory distress. The rScO_2_ values were blindly recorded to avoid modification of the ventilator settings based on such values. rScO_2_ analyses were performed only over the last 5 min of each ventilation mode to allow the newborn to stabilize after the switch between the modes. As blinded intervention with either of the two modes was impossible, we limited bias by using the rScO_2_ level as the primary outcome; this is objective and reproducible. The principal limitation of our work is that the study was ceased before we enrolled the *a priori*-calculated number of required participants due to recruitment difficulties.

## Conclusion

nHFPV was not inferior to nCPAP as determined by rScO_2_ levels when used to manage respiratory distress at birth in newborns of GA ≥ 33 weeks.

## Data Availability

All data relevant to the study are provided in the manuscript.

## Funding

This study was founded by a grant from the University Hospital of Bordeaux (Appel d’Offre Interne du CHU de Bordeaux 2013).

The English in this document has been checked by at least two professional editors, both native speakers of English. For a certificate, please see: http://www.textcheck.com/certificate/oSimss

